# Geographically-targeted COVID-19 vaccination is more equitable than age-based thresholds alone

**DOI:** 10.1101/2021.03.25.21254272

**Authors:** Elizabeth Wrigley-Field, Mathew V Kiang, Alicia R Riley, Magali Barbieri, Yea-Hung Chen, Kate A Duchowny, Ellicott C Matthay, David Van Riper, Kirrthana Jegathesan, Kirsten Bibbins-Domingo, Jonathon P Leider

## Abstract

COVID-19 mortality increases dramatically with age and is also substantially higher among Black, Indigenous, and People of Color (BIPOC) populations in the United States. These two facts introduce tradeoffs because BIPOC populations are younger than white populations. In analyses of California and Minnesota--demographically divergent states--we show that COVID vaccination schedules based solely on age benefit the older white populations at the expense of younger BIPOC populations with higher risk of death from COVID-19. We find that strategies that prioritize high-risk geographic areas for vaccination at all ages better target mortality risk than age-based strategies alone, although they do not always perform as well as direct prioritization of high-risk racial/ethnic groups.

**One-sentence summary:** Age-based COVID-19 vaccination prioritizes white people above higher-risk others; geographic prioritization improves equity.

## Introduction

Distributing COVID-19 vaccines in the United States represents one of the most significant public health challenge in a century *(1)*. National guidelines issued by the CDC in December 2020 *(2)* are consistent with the evidence that the risk of death from COVID-19 increases starkly with age *(3)*. However, the guidelines ignore evidence that the risk of exposure to and subsequent infection from SARS-CoV-2, the causative agent of COVID-19, is substantially higher for younger Black, Indigenous, and People of Color (BIPOC) *(4)*. As a result, vaccine prioritization based on age may exacerbate racial/ethnic inequities in COVID-19 burden because BIPOC populations are generally younger than the white population, more likely to be infected at younger ages, and at higher risk of dying from COVID-19 at younger ages *(4, 5)*.

In contrast, prioritizations that consider other dimensions of risk alongside age may more effectively target those at greatest risk of COVID-19 death while reducing racial and ethnic inequities. Yet not all targeted approaches are feasible in practice. While BIPOC populations have notably higher COVID-19 age-specific mortality, distributing vaccines based on race and ethnicity may not be legally viable *(6)* or politically tenable *(7–9)*. Further, a race-based approach may be perceived as discriminatory, given long-standing medical racism *(6)*. Instead, geographic targeting, using indices of health or COVID-19 mortality, may be more practical, more resistant to legal challenges, and still more equitable than strategies based on age alone *(10)*.

Here, we analyze four paired sets of alternative vaccination prioritization strategies and evaluate their sociodemographic and health equity implications. Our framework is based on maximizing the hypothetical COVID-19 mortality risk in the vaccine-eligible group using the observed COVID-19 mortality in 2020 (i.e., prior to mass vaccine rollout) as a proxy measure of risk. Given fixed vaccine supply, maximizing the mortality risk of the eligible should maximize the deaths directly averted through vaccination by directing vaccines to the people at highest risk *(11)*. In addition, maximizing the mortality risk of the eligible also improves equity in the sense that it does not prioritize lower-risk populations above higher-risk populations. Our analyses explore the intersection of this risk equity with the vaccine access of BIPOC populations and socioeconomically deprived neighborhoods. To reflect the COVID-19 mortality risk of the general population, we excluded those already prioritized in Phase 1A (long-term care residents and health care workers). We assumed policymakers and health departments aim to prioritize vaccinations for the groups with highest COVID-19 mortality risk *(11)* (rather than with highest risk of transmission *(12, 13)*), in the context of limited vaccine supply. Other COVID-19 vaccine modeling studies consider which age groups to prioritize *(14)* and various trade-offs between age, comorbidities, and occupations *(11, 13, 15–17)*. Here, we compare strategies for vaccinating the general population based on age, race and ethnicity, and alternative measures of geographic risk.

As concrete examples, we used individual-level death certificate data from California and Minnesota. These two states are socioeconomically and demographically distinct. They have experienced divergent pandemic trajectories and, according to a recent CDC analysis, differential success in vaccinating their most vulnerable residents *(18)*. We can thus compare the health equity implications of the four sets of vaccine prioritization strategies in two different populations, showing how this framework can be flexibly applied across diverse settings.

### Age-based prioritization alone results in substantial racial and ethnic disparities in averted deaths

We found that sequential age-based prioritization alone would result in substantial racial/ethnic disparities in deaths averted. For example, vaccinating all people aged 75+ would have prevented about two-thirds of white COVID-19 deaths (CA: 67%; MN: 65%). Yet, for California and Minnesota respectively, this age-based prioritization alone would have prevented only 42% and 34% of Black COVID-19 deaths, 35% and 27% of Latino COVID-19 deaths, and 63% and 32% of Asian and Asian-American COVID-19 deaths (Figure 1, top row; Figure S1). These stark differences reflect both that the white population is substantially older than most BIPOC populations and that COVID-19 mortality reaches high levels at substantially younger ages in BIPOC populations (Figure S2, top row). Age-based prioritization therefore reduces much more of the total risk in white populations compared to BIPOC populations.

**Figure 1.**
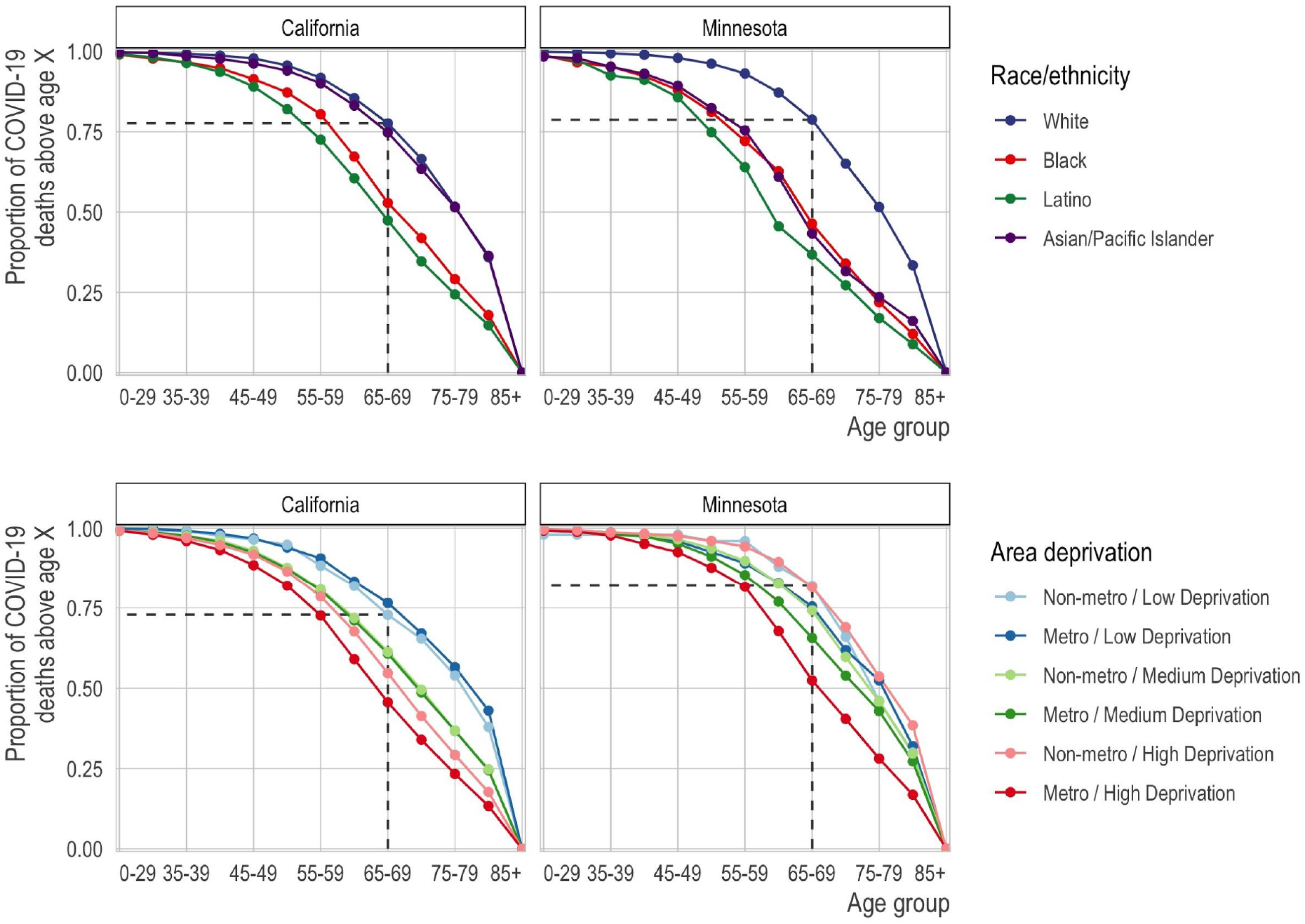
Proportion of COVID-19 deaths by race/ethnicity (top row) or geography (bottom row) and age group (x-axis) for each state (columns). Each line corresponds to the proportion of deaths (y-axis) at or above each successive age group (x-axis). In the top row, each line corresponds to a racial/ethnic category. For reference, we show the proportion of deaths among non-Hispanic whites ages 65 and older. For nearly all other racial/ethnic groups, the proportion of deaths at age 65 is lower. Correspondingly, for nearly all other racial/ethnic groups, the same proportion of deaths occurs at substantially lower ages. In the bottom row, each line represents a metropolitan area and deprivation level. Darker shades are metropolitan while lighter shades are non-metropolitan. Blue is low deprivation, green is medium deprivation, and red is high deprivation. The reference lines show the proportion of deaths at ages 65 and above among non-metropolitan, low deprivation areas.

A consequence of this multidimensional COVID-19 mortality risk is that structurally disadvantaged groups often have mortality that exceeds the state aggregate rate for age groups that are 10 or even 15 years older. For example, if mortality at ages 65-69 is sufficiently high to merit vaccine priority, the same would be true for (in California) Latinos older than 55 or (in Minnesota) BIPOC as a whole who are older than 50, because their COVID-19 mortality exceeds their state’s aggregate COVID mortality at ages 65-69 (Figure 2, top row; Figure S2).

**Figure 2.**
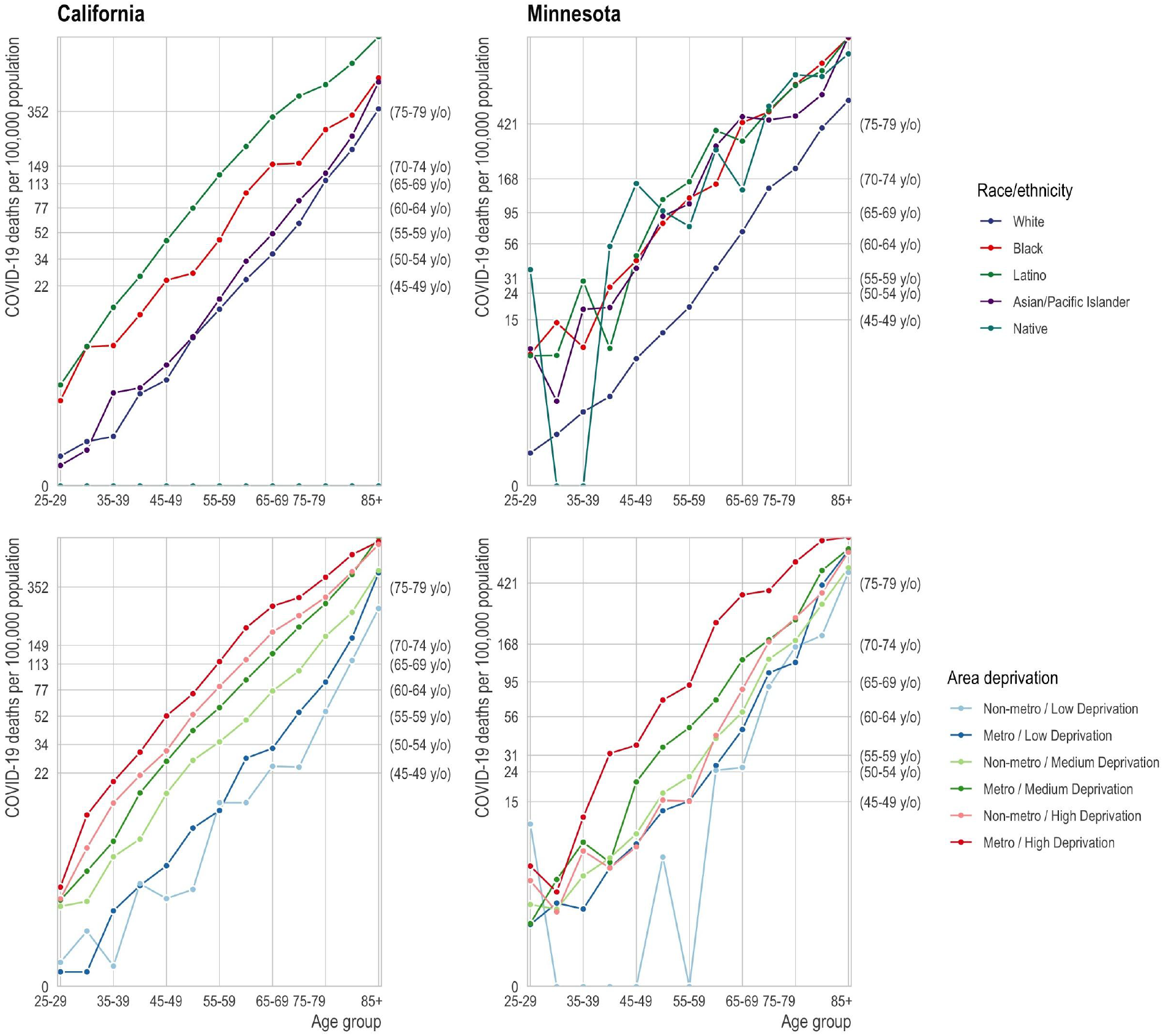
Age-specific mortality rate by race/ethnicity (top row) and geography (bottom row). Top row: The mortality rate (y-axis) by age (x-axis) varies by race/ethnicity (colors) with the non-Hispanic white population (blue) experiencing substantially lower mortality at any age relative to the BIPOC (red) and Latino (green) populations. Age-based eligibility rules ignore this variation. The secondary y-axis on the right shows the age group corresponding to the state-wide age-specific mortality rate. For example, in California, the age-specific mortality rate for non-Hispanic white 65-69 year olds is 33 per 100,000, close to the state average for 50-54 year olds (secondary y-axis) and to 45-49 year old BIPOC and Latinos. Bottom row: The mortality rate (y-axis) by age (x-axis) varies by area deprivation index (ADI; colors). We divide areas into “Metro” (lighter shades) and “Non-metro” (darker shades). We define “Metro” as the seven counties in the Twin Cities metropolitan area in Minnesota and Los Angeles, San Diego, San Francisco, Santa Clara, and Fresno counties in California. Non-metro areas include all census tracts outside of the metro category. Low deprivation is defined as an area deprivation index of 1-3, medium deprivation is 3.01-7.49, and high deprivation is 7.5-10.

In the first set of paired, alternative vaccination strategies, we compare sequential age-based vaccination (in five-year age groups) to vaccination schedules that combines the same age thresholds with race/ethnicity-age groups whose COVID-19 mortality exceeds that of the aggregate COVID mortality for the youngest eligible age group (e.g., ages 65-69 vs. ages 65-69 plus BIPOC ages 50-64 in Minnesota). We found that prioritizing vaccination for race-age groups with the highest risk would better target vaccination to high-risk individuals (Figure 3). Yet the legal, political, and practical barriers to such race-based prioritization motivates the research questions addressed in the remaining three comparison sets, which consider to what extent geographic prioritization can achieve similar ends of targeting high-risk individuals and improving racial equity in vaccination, compared to age-based rules that, in practice, prioritize white populations.

**Figure 3.**
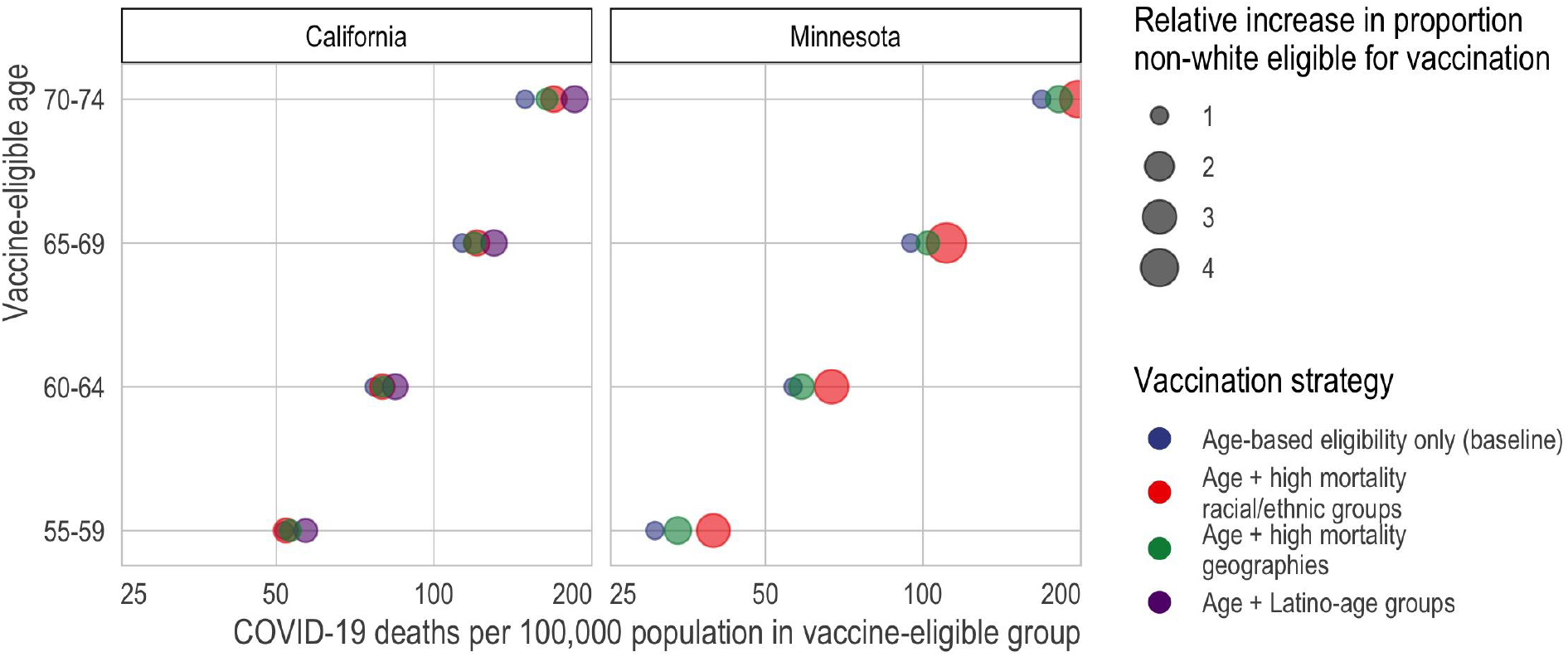
Death rate of vaccine-eligible groups under different vaccination scenarios. Here, we compare the predicted mortality rate (x-axis) of different types of vaccine allocation strategies (color) based on age (y-axis). Specifically, we compare age alone (blue), age in combination with racial/ethnic groups (red), age in combination with high mortality locations (green), and for California—the larger, more diverse state—age in combination with the highest mortality racial/ethnic group, which in California is Latinos (purple). In all cases and across all ages, incorporating additional, younger but higher risk groups improves the efficiency of the rollout and reduces racial/ethnic inequities. In California, targeting high mortality geographies (green) achieves nearly the same efficiency as prioritizing disadvantaged racial/ethnic groups as a whole (red). In Minnesota, incorporating high mortality racial/ethnic groups always outperforms incorporating high mortality geographies; however, high mortality geography still improves the alignment of vaccine allocation with COVID-19 mortality risk.

### Geographic prioritization based on area-level deprivation improves equity and averts more deaths

In the second set of alternative vaccination strategies, we compare sequential age-based vaccination to vaccination schedules that also prioritize geography-age groups whose COVID mortality exceeds that of the aggregate for the youngest eligible age group. While age-based prioritization for the 75+ age group alone would have prevented nearly two-thirds of COVID-19 deaths in advantaged neighborhoods (CA: 67%; MN: 62%), it would have prevented only 34% and 40% of COVID-19 deaths in deprived neighborhoods in major metropolitan areas in California and Minnesota, respectively (Figure 3, Figure S3).

Compared to age-based prioritization alone, prioritizing by area-level deprivation can better target high-risk groups (Figure 3, Table S1). In California, geographic prioritization targets mortality about as effectively as prioritizing BIPOC as a whole, although not as well as prioritizing Latinos (the highest-risk racial group) specifically; in Minnesota, geographic prioritization is less effective than prioritizing BIPOC populations. Geographic prioritization also increases racial equity in Minnesota but does so only very modestly in California.

### Universal adult vaccination in the highest-mortality neighborhoods can improve equity and avert more deaths

In the third comparison set, an alternative geographic prioritization strategy would directly identify Census tracts with historically higher COVID-19 mortality rather than proxying risk by area deprivation and major metropolitan status. This strategy mirrors one adopted by some states *(19)*. Compared to statewide sequential age-based prioritization alone, adding vaccination for all adults (ages 20+) in the highest mortality tracts would generally improve the targeting of high-mortality groups in contexts where it also improves vaccine uptake among older people in the high-mortality tracts, but not in contexts where vaccinating the high-mortality tracts adds vaccination only for the youngest (not among those who were already eligible due to their age) (Figure 4; see details in Materials and Methods). Prioritizing high-mortality tracts would also dramatically increase vaccine access for BIPOC communities (Figure 5). These results are qualitatively robust to a sensitivity analysis that assumes that a large portion of “high-mortality tracts” included unidentified long-term care facilities whose deaths should be excluded from the analysis (Figure S4; see details in Materials and Methods).

**Figure 4.**
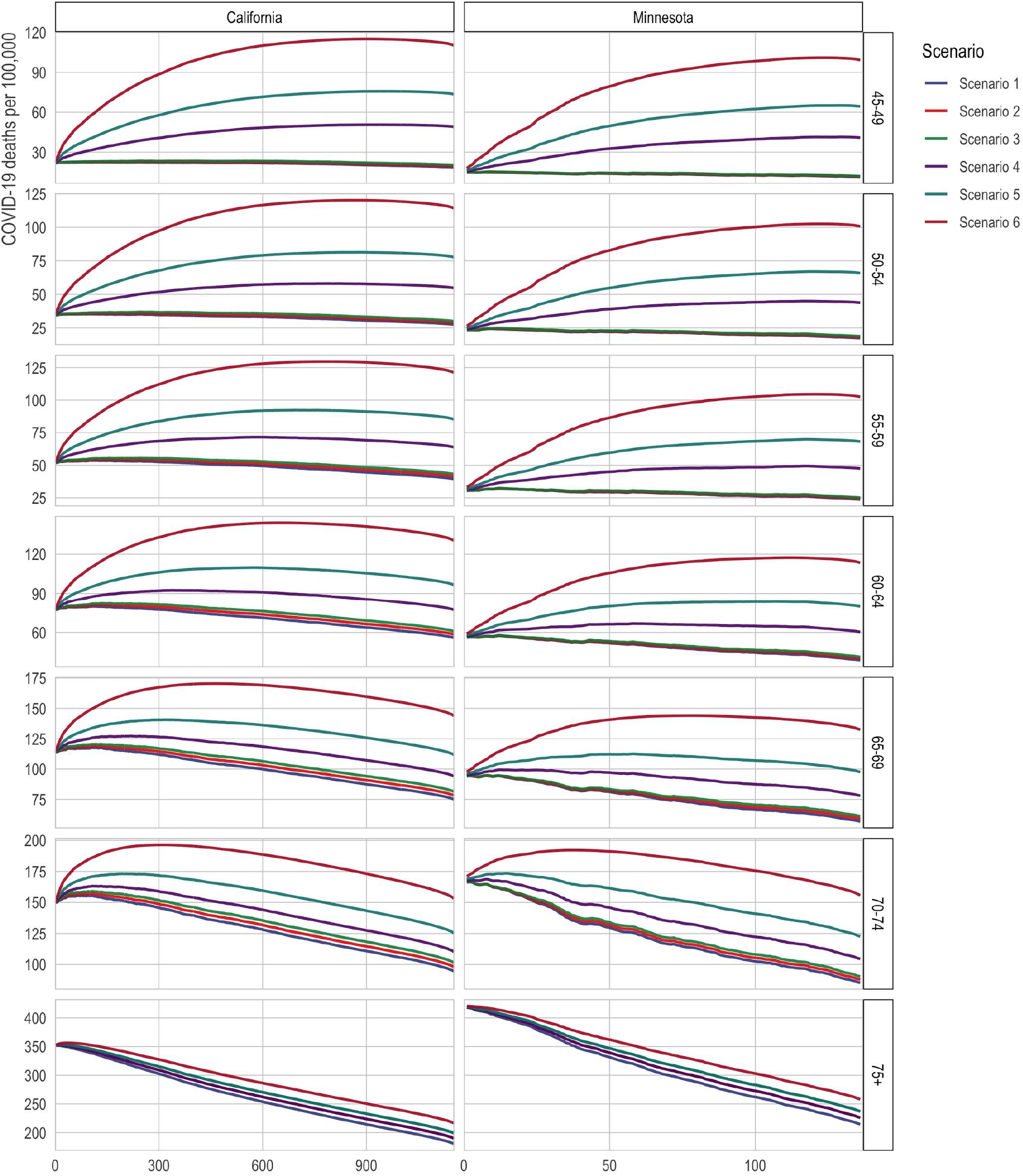
Death rates among the eligible with direct targeting of high-mortality Census tracts. The x-axis is the number of tracts in which all adults (ages 20+) are prioritized for vaccination. The y-axis is 2020 COVID-19 deaths per 100,000; a higher death rate among the eligible indicates better targeting of vaccines toward high-risk individuals. The lines correspond to alternative scenarios as described in the text of the Materials and Methods.

**Figure 5.**
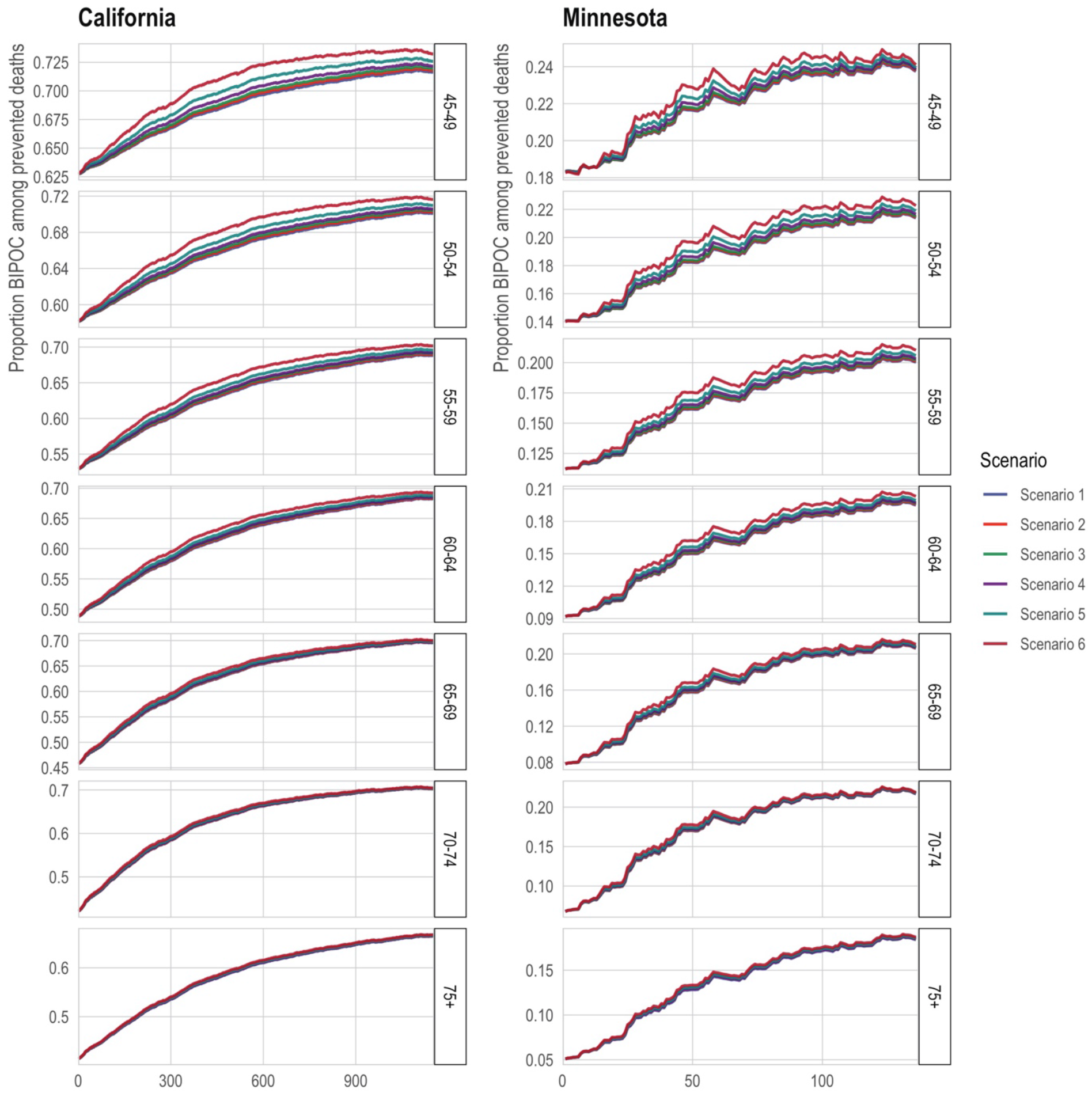
Proportion non-white among the eligible with direct targeting of high-mortality Census tracts. The x-axis is the number of tracts in which all adults (ages 20+) are prioritized for vaccination. The y-axis is the proportion of the state’s eligible population that is non-white. The lines correspond to alternative scenarios as described in the text of the Materials and Methods.

For illustration, in California, if prioritizing tracts does not increase vaccine uptake among the oldest tract residents (who would already be eligible by age), then vaccinating the 500 highest-mortality tracts would decrease the mortality averted by 9% compared to vaccinating the 65-69-year-olds alone. (The inflection point, where prioritizing all adults in a tract is neutral, occurs at around 250 tracts under the assumption of no improved older-age vaccination.) However, if prioritizing tracts increases vaccine uptake by 50% among the oldest, already-eligible residents of those tracts, then vaccinating the 500 highest-mortality tracts would increase the averted mortality by 22%.

### Universally lowering the age of eligibility averts fewer deaths and is less equitable than selectively lowering eligibility age

In the fourth comparison, we consider alternative strategies aimed at increasing racial equity in vaccination: substantially lowering age thresholds across the board, as some states have adopted with this motivation *(20)*, versus selectively lowering age thresholds for high-mortality geographies. We compare these strategies at two critical junctures representing “early” and “late” vaccine rollout points: when vaccinating the 70-74 age group and when vaccinating the 55-59 age group (Figure 6; see details in Materials and Methods). The benefits of selectively lowering the age threshold, for maximizing the extent to which eligibility aligns with those at highest mortality risk, are substantial: for the older ages, selective lowering better targets the aggregate mortality risk of the eligible by 55% (159 vs. 103 deaths per 100,000) in California, and 88% (178 vs. 95 deaths per 100,000) in Minnesota; for the younger ages, selective lowering better targets mortality risk among the eligible by 51% (52 vs. 34 deaths per 100,000) in California, and 40% (32 vs. 23 deaths per 100,000) in Minnesota. However, in California, selective lowering of the age threshold does not meaningfully increase the proportion of vaccine-eligible people who are BIPOC for either early or late rollout. For Minnesota, it increases the proportion of vaccine-eligible who are BIPOC modestly (11% vs. 8% for the older ages; 18% vs. 14% for the younger ages).

**Figure 6.**
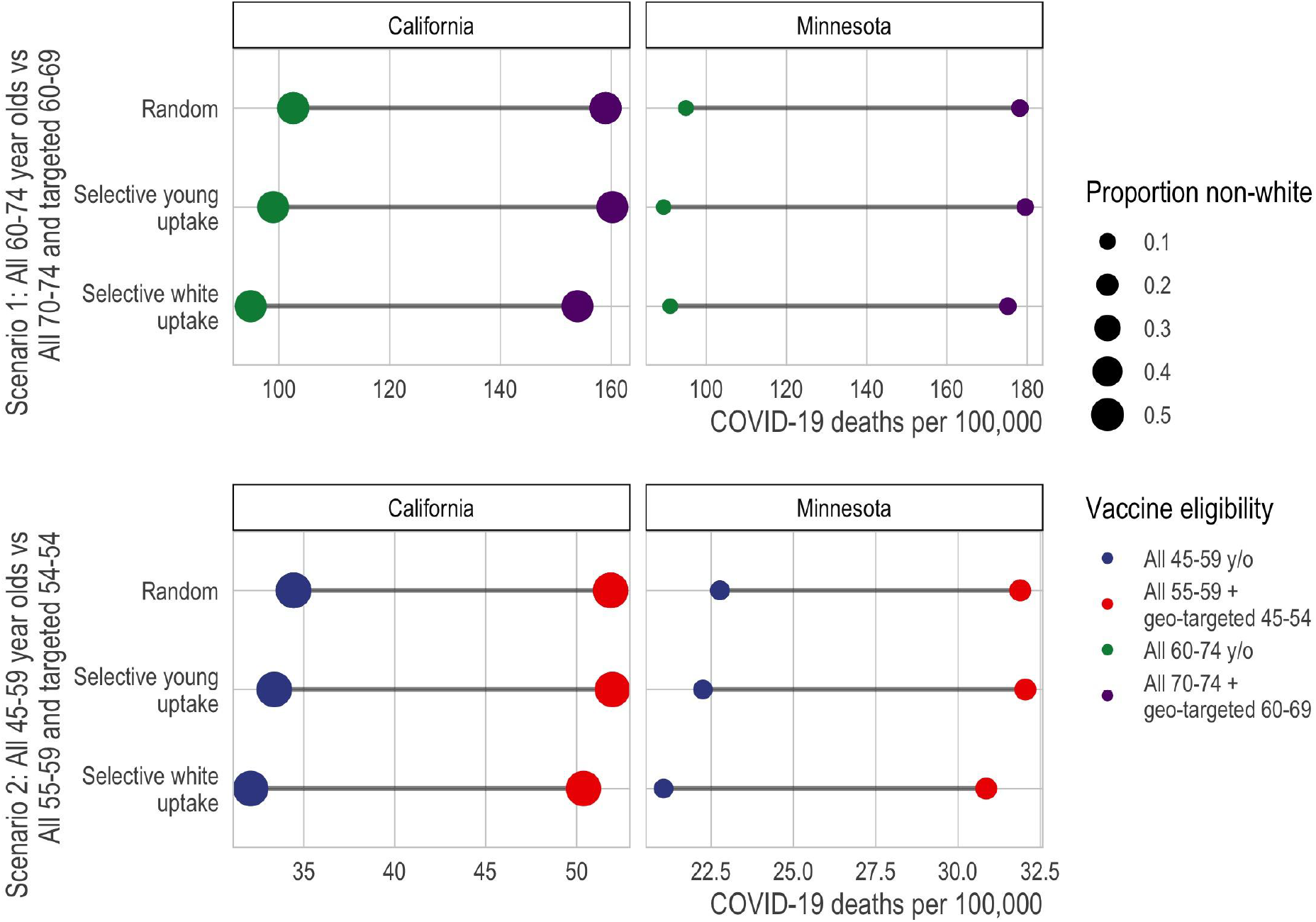
Death rate of vaccine-eligible populations under alternative strategies designed to increase equity for BIPOC populations. Here, we compare the targeted mortality rate (x-axis) of different types of vaccine allocation strategies (color) under alternative assumptions about vaccine uptake (y-axis). Specifically, we compare strategies that universally reduce the age at eligibility (blue) to strategies that retain a higher age at eligibility but drop to a much younger age for high-risk geographic units, defined by Area Deprivation Index and major metropolitan status. In each panel, the first line assumes that vaccine uptake is random among the eligible; the second assumes that, conditional on eligibility, each successively younger 5-year age group increases uptake by 10% gr if the age threshold is high and by 50% if the age threshold is low; and the third line assumes that, conditional on eligibility, whites experience a 10% greater uptake if the age threshold is high and 50% if the age threshold is low. The second and third lines indicate that a larger eligible group relative to vaccine supply may exacerbate selective uptake of lower-risk eligible people.

An additional shortcoming of broadly lowering age thresholds is obscured by the assumption of random uptake among the eligible: broadly lowering the age threshold can exacerbate the selective uptake of lower-risk individuals to the extent that the size of the eligible group exceeds the available vaccine supply. To capture this phenomenon, we compare the mortality risk among the vaccinated, and proportion BIPOC among the vaccinated, under varying degrees of selective uptake among whites and selective uptake among younger eligible people. We find that, to the extent that creating a larger eligible population might exacerbate selective uptake by badly outstripping vaccine supply (e.g., white people being 25% vs. only 10% more likely than BIPOC people to access vaccines when eligible, the former number in line with observed rates *(21)*), geographic targeting will be even more effective at targeting high-risk groups and will also produce more equitable vaccination (Figure 6). At these relatively low rates of selective uptake, the difference made by selective uptake is small relative to the differences made by the vaccination schedules even assuming random uptake. Larger rates of selective uptake produce more dramatic divergences between the schedules (Figure S5).

## Discussion

Our results showed, first, that strict age-based vaccination strategies disproportionately benefit the white population. For example, in both California and Minnesota, after excluding long-term care populations and health care workers, more than three-quarters of white COVID-19 deaths occurred above age 75, but half or fewer of Black and Latino deaths. This prioritization might be justifiable if older populations were at higher risk than younger populations, irrespective of race, much as prioritizing residents of long-term care facilities in Phase 1a resulted in prioritizing a largely white population at overwhelming risk *(22)*. However, we show that this justification does not apply to age-based vaccination after Phase 1a. For example, when state vaccination eligibility is extended from 75+ to 65+, the mortality rate among the newly eligible is lower than the mortality rate among BIPOC groups that are 10 or 15 years younger yet still ineligible for vaccination. These age-ineligible, yet high-risk, BIPOC groups may have to wait for up to three months longer to be eligible for vaccination *(23)*. These results underscore the implications of prioritizing vaccine allocation based on the 65+ age threshold, as many states implemented in January 2021.

Second, compared to a vaccine eligibility strategy based on age alone, a strategy that combines geographic location based on socioeconomic characteristics with age-based eligibility--such as by extending eligibility to the geographic and age groups with higher mortality than the youngest age-eligible group--better aligns with risk of COVID-19 mortality. The total improvements in risk coverage from this age-geography prioritization are fairly modest (improving the targeting of high-mortality groups by 3-10% across age groups and states) because the populations added through geographic prioritization are small relative to the five-year age groups in each state, so they have only a relatively small effect on aggregate risk among the eligible. However, the small size of the populations that would additionally become eligible also implies that geographic prioritization has a low direct opportunity cost, as only a small number of vaccines need to be allocated to high-risk geographies to achieve the equity gains of targeting.

Third, in the context of vaccine scarcity, efforts to save the most lives possible and to save lives equitably can be at odds *(10, 24)*. Our results suggest that, in some cases, directing vaccination efforts at small, high-risk geographic areas without regard to age can improve on efforts to target older ages throughout the state, especially when such geographically targeted efforts improve vaccine uptake among older residents of high-risk areas. These results suggest that states should consider targeting broad swaths of the population (e.g., all adults) in highly specific geographic contexts when--and, from the perspective of directly reducing mortality, perhaps only when--this targeting allows for tactics that allow older residents to be more effectively reached. Such tactics could include home visits *(25, 26)*, walk-in pop-up clinics *(27)*, assigning appointment slots to all residents *(28)*, and other forms of direct outreach. Such approaches may be especially likely to succeed in increasing uptake among the highest-risk when high-risk populations are vaccine-hesitant but might be more likely to adopt vaccination as others in their networks become vaccinated, and to the extent that such approaches increase framing of vaccination as the local default *(28)*. Such direct outreach might be an effective strategy to vaccinate very high-risk populations quickly.

Moreover, broadly prioritizing all adults in the highest-mortality neighborhoods may be even more effective than the results here suggest. To the extent that groups with disproportionately high mortality also have disproportionate incidence of infection, the mortality-based results here may understate the benefits of better targeting at-risk groups. Because people live in segregated communities, people at heightened risk of COVID-19 death are likely to interact with others at elevated risk. Thus, prioritizing vaccination more effectively by neighborhood can potentially have multiplier effects as vaccinating relatively old residents reduces mortality directly and vaccinating younger residents reduces transmission to high-risk older people *(29)*.

Fourth, several states have recently extended age eligibility to age 50+ *(30)* and even to all adults *(31, 32)*, with reductions in the age at eligibility sometimes driven by a recognition that BIPOC people die of COVID-19 at younger ages on average *(33)*. However, large universal drops in the age threshold for eligibility have the consequence of targeting risk quite poorly. We show that, compared to such a strategy, an alternative strategy that incorporates only high-risk geographies at younger ages does substantially better at prioritizing people with higher mortality risk. This is especially true in the context of disproportionate vaccine uptake by the advantaged among the eligible. However, our vaccine uptake simulation results suggest that small to moderate rates of selective uptake make relatively little difference in the extent to which each vaccination strategy succeeds in prioritizing high risk people, compared to the large difference made by the choice of strategy itself.

Our results additionally suggest that better-optimized vaccination strategies should consider local demographics, intersectional risks, and both large-scale (e.g., large metro areas) and small-scale (e.g., Census tract disadvantage) geographic stratification. For example, in both states, disadvantaged metropolitan Census tracts had distinctly higher COVID-19 mortality than all other geographies. Yet we found that geographic risk was more stratified by area deprivation index in California and more stratified by major Metro status in Minnesota, implying that a one-size fits all approach may be sub-optimal given vast demographic and geographic heterogeneity across states. Our results underscore the need for each state to individually consider what metrics would be most impactful for vaccine prioritization that both simultaneously maximizes the reduction in deaths due to COVID-19 while also ensuring a fair and equitable approach.

This study has several limitations. First, the calculations reported in this analysis are based on mortality data obtained from January to December 2020. Therefore, to the extent that mortality patterns by age, race/ethnicity, and place have changed over the course of the pandemic (e.g., responses to selective shutdowns or social distancing patterns), our results may not reflect future deaths averted by vaccination. Second, we were only able to evaluate strategies that prioritize based on information included in death certificates, which notably excludes strategies based on comorbidities. Third, in some of our analyses of racial equity, we included all Black, Indigenous, and People of Color (BIPOC) into one racial/ethnic category. Collapsing across diverse racial/ethnic and Indigenous populations poses challenges with respect to generalizability and implies a universal lived experience which does not exist *(34, 35)*. However, combining groups enabled us to make direct comparisons between states (including a smaller, predominantly white state, Minnesota). Fourth, our study focused on vaccine eligibility and considered vaccine access only via selective uptake simulations. Yet access given eligibility may be as important as eligibility per se in determining equitability in COVID-19 vaccination. Moreover, some strategies are easier to implement than others. Geographic prioritization strategies require states to leverage data to determine where to target, whether broad indexes of risk like the area deprivation index or direct measures of where deaths have been concentrated in the state. Strategies that prioritize active outreach in small, high-risk areas require coordination and other resources and, to be effective, may require staff with linguistic competence and community connections that health departments may lack. Finally, vaccination strategies that are not widely perceived as legitimate can undermine social solidarity and increase efforts to flout the rules *(36)*, and we did not evaluate whether geographic prioritization is likely to be widely perceived--or can be made to be widely perceived--as fair.

A central argument for age-based vaccination schedules is that they may minimize administrative burdens that may undermine more targeted schedules by preventing the eligible people who are at highest risk from accessing the vaccine. For example, targeting comorbidities may inadvertently exclude people without primary care doctors *(37)*. Geographic prioritization strategies, like those explored here, may chart a middle path between, on the one hand, broad eligibility criteria that minimize administrative burden and, on the other, highly-targeted criteria that aim to direct vaccines at groups with the highest mortality risk. Geographic prioritization is not free of administrative burden, particularly for those without secure housing, who need to be reached with alternative strategies. And in particular, since few individuals know their Census tract, the prioritization strategies considered here would require individuals to check the eligibility of their addresses (e.g., through an online system or over the phone) or to be proactively contacted by state health systems; merely placing vaccination sites in high-risk neighborhoods does little to ensure that residents of those neighborhoods will be the people vaccinated *(38)*. In many spheres of service provision, there are strong arguments in favor of universalist systems that minimize the burdens of demonstrating eligibility *(39)*. Yet the vaccine rollout is a unique context in which the supply has been inflexibly scarce, making a truly universal approach untenable. Given this, strategies that prioritize residents of the neighborhoods where risk of dying of COVID-19 has been heavily concentrated could protect people whom age-based strategies exclude, in spite of their heightened risk of death.

## Data Availability

All data needed to evaluate the conclusions in the paper are present in the paper and/or the Supplementary Materials. The data can be downloaded from a permanent repository at https://osf.io/tzeq3/?view_only=191ff8cfbd104e3aa2e76518af554943

## Acknowledgements

We thank Elaine Hernandez, Maria Glymour, Michelle Niemann, Govind Persad, and Matthew Plummer for helpful discussion. We thank the California Department of Public Health and Minnesota Department of Health for access to death certificate data.

## Author contributions

EWF and MVK conceived and designed the study. EWF, MVK, Y-HC, MB, DVR, KPL, ARR, KBD, JPL acquired, analyzed, and interpreted the data. EWF, MVK, ARR, ECM, MB, KAD, JPL drafted the initial version of the manuscript. All authors provided critical revisions. All authors approve of the final manuscript and take final responsibility for the decision to submit for publication.

EWF and MVK had direct access to and verified the underlying data. In addition, JPL had full access to the Minnesota death certificate data and Y-HC had full access to the California death certificate data.

## Data availability statement

In accordance with our data use agreement, individual-level data are not publicly available. When possible, we will provide aggregated data upon request and in accordance with our data use agreements.

## Supplemental Materials for

### Material and Methods

#### Mortality data

We used death certificate data provided by the California and Minnesota Departments of Public Health to identify all deaths due to COVID-19 from January 1, 2020 to December 31, 2020 (N=21,668 for California; N=5,803 for Minnesota). We exclude 2021 deaths to limit distortion from vaccinations, since our goal is to estimate the mortality risk of various groups were they unvaccinated. For Minnesota, deaths are considered to be COVID-19 deaths if ICD code U07.1 appears in any cause of death line on the death certificate. For California, deaths are considered to be COVID-19 deaths if ICD code U07.1 appears in the underlying cause of death line, which excludes 1,565 deaths with COVID-19 as a contributory cause. To reflect the underlying COVID-19 mortality risk that would be observed in the general population, we excluded decedents who would be eligible for Phase 1A of the vaccine. Specifically, we removed COVID-19 deaths that occurred in long-term care facilities or nursing home residents (N = 3,114 for California; N = 3,070 for Minnesota). Deaths that occurred in long-term care facilities were identified in California by, first, using death certificate place of death and, second, matching the location of deaths to a comprehensive list of long-term care facilities. In Minnesota, these deaths were identified by the death certificate (see below for details and sensitivity analyses). In addition, we removed deaths among health care workers in California (N = 759); however, we were unable to remove health care worker deaths in Minnesota (N = 28).

We limited analyses of specific racial groups to non-Hispanic white, non-Hispanic Black, Latino, and Asian/Asian-American populations because they are the largest populations in both states and we have greater confidence that death certificate racial assignments match population racial categorizations. Latino or Hispanic identity took precedence over racial group assignment. In Minnesota numerators and in both state denominators, the Asian group includes Pacific Islanders; in California, those are coded as “other-race” deaths (N=503), resulting in a small undercount of the COVID-19 death rates for Asians in California. We treat all non-white populations (including those recorded as “Other race” on death certificates) as the BIPOC group.

#### Population data

Official 2020 population estimates are not yet available. We projected 2020 population estimates by race/ethnicity and age using historical population counts. Specifically, we used the Census Bureau July 1st population estimates by race, sex, and single-year of age for 2019 and the number of deaths by race, sex, birth cohort and age that took place between July 1, 2019 and June 30, 2020 in each state. Assuming zero net migration for each state during this 12-month period, we implemented the cohort-component method for each sex and race to estimate the July 1st population estimates by race, sex, and single-year of age for 2020. These calculations were performed after redistributing deaths of “Other” races, including multiple races (i.e. other than “Non-Hispanic White”, “Non-Hispanic Black”, “Non-Hispanic Native American”, “Non-Hispanic Asian”, “Hispanic”) proportionately over the other races for each sex and within each age group. Similarly, we redistributed deaths of unknown ages proportionately over all known ages within each race, sex, and age group category. We verified that our population estimates for July 1, 2020 were consistent with past trends in the state (using Census Bureau July 1 population estimates for 2010 through 2019) *(1)* for each combination of race, sex, and 5-year age group.

In order to estimate Census tract-specific populations for each race-, sex-, and age-specific group, we used the 2013-2018 National Historical Geographic Information System (NHGIS) estimates *(2)*, which are the most recent available, as a baseline measure. The NHGIS estimates are in 10-year age bands. To produce 5-year age bands, we used the single-year-age projections of the 2020 population to estimate the proportion of each race- and sex-specific 10-year age interval that is in the older or younger 5-year group in that interval. Finally, we scaled the resulting Census tract-specific estimates up to the projected 2020 population size using the ratio of the 2020 population to the 2013-2018 NHGIS estimates for each race-, sex-, and 5-year-age-specific population. This procedure assumes that differential population growth across geographic areas in each state is proxied by the resident demographics.

#### Geographic disadvantage

We define geographic disadvantage using the area deprivation index (ADI) and metropolitan status. For each census block group, the ADI provides a score ranging from 1 (low deprivation) to 10 (high deprivation) based on 17 area-level measures about education, employment, housing quality, and poverty *(3)*; the scores represent deciles of the state distribution of multidimensional deprivation. We use the ADI rather than the widely-used Social Vulnerability Index (SVI) because the SVI includes the racial composition of geographic areas as a component of vulnerability *(4)*, whereas our goal is to evaluate facially race-neutral geographic targeting. For each Census tract, we took the population-weighted mean ADI score and categorized tracts as low deprivation (<=3), medium deprivation (3.01 to 7.49), and high deprivation (>=7.5). The asymmetry in the cutpoints for advantage and disadvantage reflects that there are extremely few COVID-19 deaths in areas with ADI<2.5 in Minnesota, producing unstable age-specific mortality rates without including slightly less advantaged tracts. In addition, we categorized tracts by metropolitan status. In Minnesota, metropolitan tracts were the seven counties in the Twin Cities metropolitan area (collectively representing about 56% of the state population). In California, metropolitan tracts were those in Los Angeles, San Diego, San Francisco, Santa Clara, and Fresno counties (collectively representing about 44% of the state population). Non-metro areas include all Census tracts outside of the metro categorization.

#### Statistical analysis

We used COVID-19 mortality in 2020 to proxy COVID-19 mortality risk in 2021 in the absence of vaccination, thus ignoring changes from selective mortality, improved treatment, and evolving patterns of risk. We do not assume that risk in 2021 in the absence of vaccination would equal risk in 2020, but we do assume that, across groups, 2021 risk in the absence of vaccination would be proportional to 2020 risk. This assumption allows us to compare the aggregate risk of the eligible under various vaccination strategies.

Our main results assume that vaccine uptake is random among the eligible group. However, we also compare prioritization schedules in the context of differential vaccine uptake among the eligible by race or by age (to varying degrees), reflecting that the available vaccine supply exceeds demand among the eligible. These simulations are described below.

This study was deemed exempt from full review by the University of Minnesota institutional review board (STUDY00012527) and was approved by the California Health and Human Services institutional review board (Project number: 2020-109).

### Census tract estimates (Comparison set 3)

To assess schedules that include high-mortality Census tracts directly, we rank tracts in each state by their COVID-19 mortality. This analysis, like our other geographic analyses, is limited to deaths that could be successfully geocoded (excluded deaths are N=58 in California; N=165 in Minnesota). We limit the tract ranking to tracts with at least 1,000 residents and at least 5 COVID-19 deaths (which means that the ranking is limited to relatively high-mortality tracts; N=1,089 tracts in California; N=114 tracts in Minnesota) in order to reduce the inclusion of tracts that had high mortality for idiosyncratic reasons in 2020 that would not have applied in 2021. A remaining danger is that some included tracts may contain unidentified long-term care facilities, which would mean that the true inflection point (at which prioritizing all adults in high-mortality tracts flips from improving to worsening mortality targeting) would occur at a lower number of tracts than our analysis suggested. This possibility is explored in a sensitivity analysis described below.

We consider tract inclusion under six scenarios, ranked by the extent to which vaccinating all adults in the tract would prioritize high-risk individuals:

1. “No tract benefit for older people”: Assumes that older people in prioritized tracts, who are eligible by age as well as tract, would be vaccinated at the same rate as their age group in the rest of the state and gain no additional likelihood of vaccination from tract priority;
2. “Tract benefit only for youngest old (smaller benefit)”: Assumes that the youngest older people in prioritized tracts, who have just recently become eligible by age as well as tract, would become 25% more likely to be vaccinated when their tract is prioritized, but older people who have been eligible longer would be vaccinated at the same rate as their age group in the rest of the state and gain no additional likelihood of vaccination from tract priority;
3. “Tract benefit only for youngest old (larger benefit)”: Assumes that the youngest older people in prioritized tracts, who have just recently become eligible by age as well as tract, would become 50% more likely to be vaccinated when their tract is prioritized, but older people who have been eligible longer would be vaccinated at the same rate as their age group in the rest of the state and gain no additional likelihood of vaccination from tract priority;
4. “Tract benefit for all older people (smaller benefit)”: Assumes that all older people in prioritized tracts, who are eligible by age as well as tract, would become 25 more likely to be vaccinated when their tract is prioritized;
5. “Tract benefit for all older people (larger benefit)”: Assumes that all older people in prioritized tracts, who are eligible by age as well as tract, would become 50 more likely to be vaccinated when their tract is prioritized;
6. “Tract benefit regardless of age-eligibility”: Assumes that all older people in prioritized tracts, who are eligible by age as well as tract, would become twice as likely to be vaccinated when their tract is prioritized (thus gaining the same absolute increase as younger people in the tract).

All six scenarios additionally assume that people in prioritized tracts who are too young to be otherwise eligible become vaccinated at the same rate as age-eligible people outside prioritized tracts.

#### Analysis of alternative equity strategies: substantially lowering the age threshold universally vs. selectively (Comparison set 4)

In these analyses, we compared the strategy of substantially lowering the age threshold for eligibility universally to the strategy of keeping the age threshold at a higher level but selectively lowering it for high-risk geography-age groups. High-risk geography-age groups are defined in the same way as in comparison set 2: geographies are stratified by area deprivation and metropolitan status, and geography-age groups are declared eligible when their mortality exceeds that of the state aggregate for the youngest eligible age group.

In these analyses, we compared these strategies at two points:

1. Scenario 1/Early vaccination: Setting statewide eligibility at age 60 vs. setting statewide eligibility at age 70 while additionally prioritizing high-risk geographies as young as 60.
2. Scenario 2/Late vaccination: Setting statewide eligibility at age 45 vs. setting statewide eligibility at 55 while additionally prioritizing high-risk geographies as young as 45 (California) or 40 (Minnesota).

We selected these age targets in order to match comparisons across states at “early” and “late” vaccination ages. In both California and Minnesota, the statewide 70-74 age group’s mortality is exceeded by residents of deprived Metro areas beginning at age 60. (In California, the 70-74 group’s mortality is also exceeded by residents of deprived non-Metro areas beginning at age 65.) In California, the statewide 55-59 age group’s mortality is exceeded by residents of deprived Metro areas beginning at age 45; in Minnesota, this age group’s mortality is exceeded by residents of deprived Metro areas beginning at age 40. (The 55-59 group’s mortality is also exceeded by residents of deprived non-Metro areas in California, and medium-deprivation Metro areas in Minnesota, beginning at age 50.) Thus, Scenario 2’s comparison statewide threshold of age 45 is conservative for Minnesota in the sense that a schedule aiming to include the most deprived neighborhoods would need to incorporate age 40, not just 45.

In the baseline analysis of these scenarios, we assumed that vaccine uptake was random among the eligible. In the selective uptake simulations (described more fully in the next section), we considered what would happen if dramatically increasing the size of the eligible group (e.g., by dropping the statewide age threshold by 10 years of age) increased the selective uptake of white people, or of younger people, among the eligible from 10% to 25%. These numbers were chosen to be roughly calibrated against data suggesting about 25% selective uptake among white people six weeks into eligibility at 65+ *(5)*.

#### Sensitivity analysis: Selective uptake

We considered results under selective uptake by white people or young people. In the race analyses, we allowed white people to be more likely, given eligibility, to access vaccines by 0% (baseline), 5%, 10%, 15%, 25%, 50%, 100%, or 150%. In the age analyses, we allowed each successively younger age group to be more likely, given eligibility, to access vaccines by the same percentages.

We operationalized selective uptake by over-weighting the relevant (white or young) populations by the appropriate amount in estimating the mortality and proportion BIPOC of the vaccine-prioritized group.

#### Sensitivity analysis: Alternative classification of long-term care deaths in Minnesota

Minnesota is unusual in that a majority of COVID-19 deaths in the state occurred in long-term care facilities. For this reason, we analyze whether results about high-mortality Census tracts could be partially driven by misclassifying some long-term care deaths as general population deaths in Minnesota. The primary identification of long-term care deaths in Minnesota is based on two death certificate fields that identify whether the death occurred in a long-term care facility and whether the decedent was a resident of long-term care. We count both as LTC deaths (so that we include, for example, LTC residents who were hospitalized shortly before their deaths). This strategy produces an undercount of LTC deaths relative to numbers estimated by the Minnesota Department of Health (MDH) through the end of 2020 (3,070 vs. 3,803).

As an alternative strategy constructed as a sensitivity analysis, we also classify as LTC deaths any deaths that are estimated to have occurred in the same location as any LTC deaths (to any cause and in any year, 2017-2021 to date), in addition to those classified as LTC deaths based on their death certificates. We count deaths as occurring in the same location if they occurred at a known address or street point within a 50-meter radius of one another.

This strategy overestimates LTC deaths, producing an estimate of 3,902. The overestimation likely occurs for two reasons. First, many establishments combine a LTC facility with independent living facilities. The MDH classifications exclude those in independent living from LTC counts and, for our purposes, this exclusion is correct because independent living residents were usually not prioritized for vaccination along with LTC residents in Phase 1a (per communication with MDH). However, our location-based sensitivity analysis will count the independent living deaths as LTC deaths as long as they occurred within 50 meters. Spot-checking locations in which large numbers of deaths were tagged as LTC and large numbers also were not tagged as LTC, on death certificates, reveals that these are, as expected, institutions that combine memory care facilities with independent living facilities. Second, this location-based classification may erroneously include residents of large non-LTC residential facilities in any individual is tagged on the death certificate as an LTC death even though their address is not an LTC facility. For example, spot checking revealed a public housing building in Saint Paul in which four deaths were “LTC deaths” but had a residential address in that building, even though the building does not contain LTC units. These deaths may be people who moved into LTC shortly before their deaths and had their longer-term address listed on the death certificate (or may have some other source of data error), but this location-based sensitivity analysis then treats *all* residents of this public housing building as LTC deaths. Because of these errors of commission, which will differentially exclude people living in high-risk housing or among high-risk individuals, the alternative strategy for identifying LTC deaths is not preferred, but is included as a sensitivity analysis because it errs in a different direction from the main analysis.

As expected, the analysis using these alternative exclusion criteria finds smaller benefits to universally vaccinating all adults in high-mortality tracts than does the main analysis (Figure S4). For example, using these criteria, we find, at best, only minimal benefit, and more often detriment, to universal vaccination at any point when prioritizing high-mortality tracts is assumed to yield no increase in vaccine uptake among older people in those tracts. When tract prioritization is assumed to increase vaccine uptake among older people by 50%, this alternative analysis finds minimal benefit or detriment to tract prioritization at the stage when 70+ ages are otherwise being vaccinated, but meaningful benefit as ages younger than 70 become eligible. Perhaps the biggest difference from the main results shown in Figure 4 and Figure 5 is that, given the very sensitive alternative measure of LTC deaths, a notably smaller number of Census tracts is considered to have sufficient population and non-LTC COVID-19 deaths to generate reliable evidence of high mortality (N=46 vs. N=136). These alternative results can be interpreted as a lower bound on the benefits of universal prioritization of high-mortality tracts in Minnesota.

## Supplemental Figures

**Figure S1.**
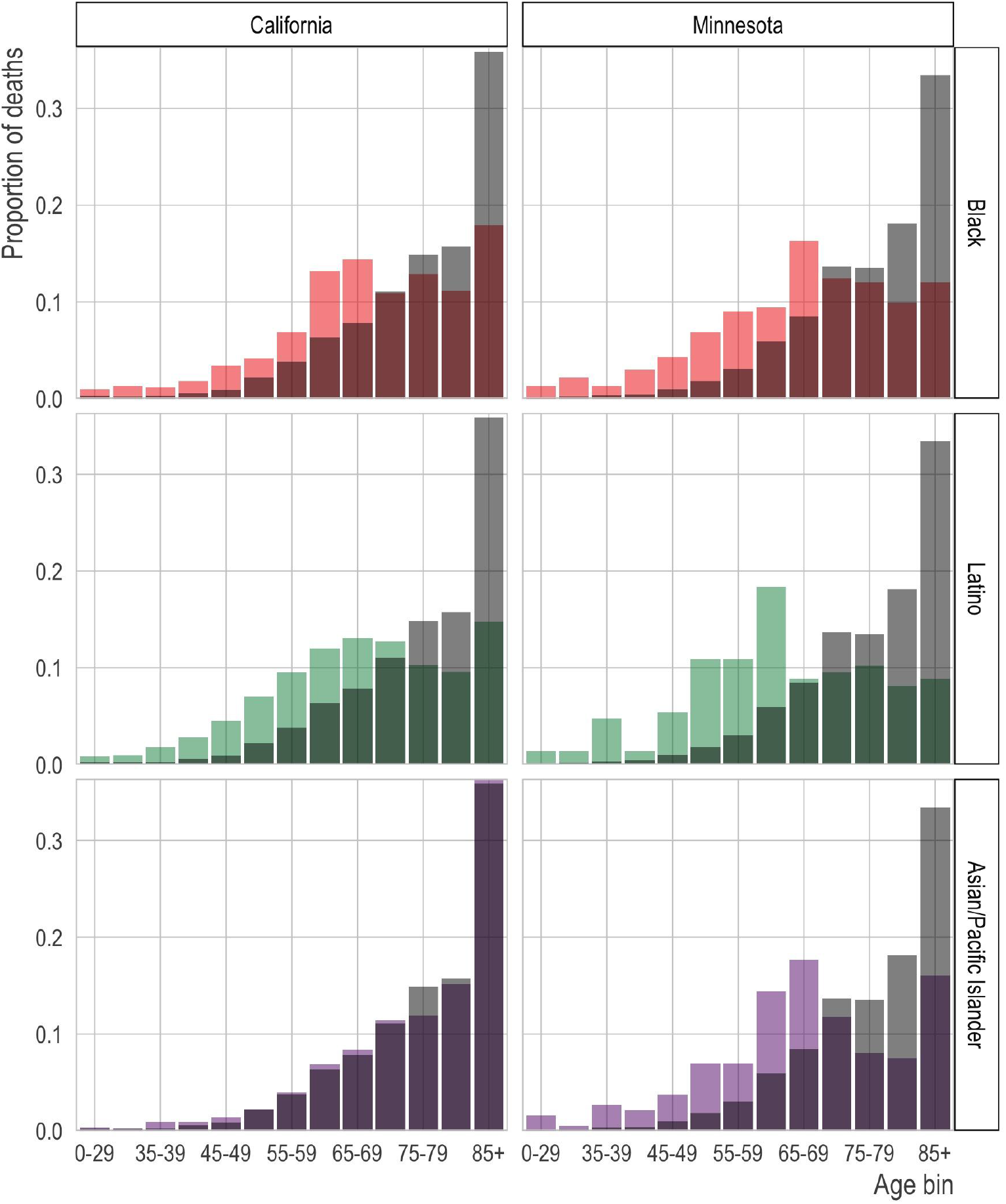
Age distribution of COVID-19 deaths by race/ethnicity and state. We show the proportion of COVID-19 deaths (y-axis) in each age bin (x-axis) by race/ethnicity (rows) for California (left column) and Minnesota (right column). The grey bars in the background of each panel show the age distribution of deaths in the non-Hispanic white population for comparison.

**Figure S2.**
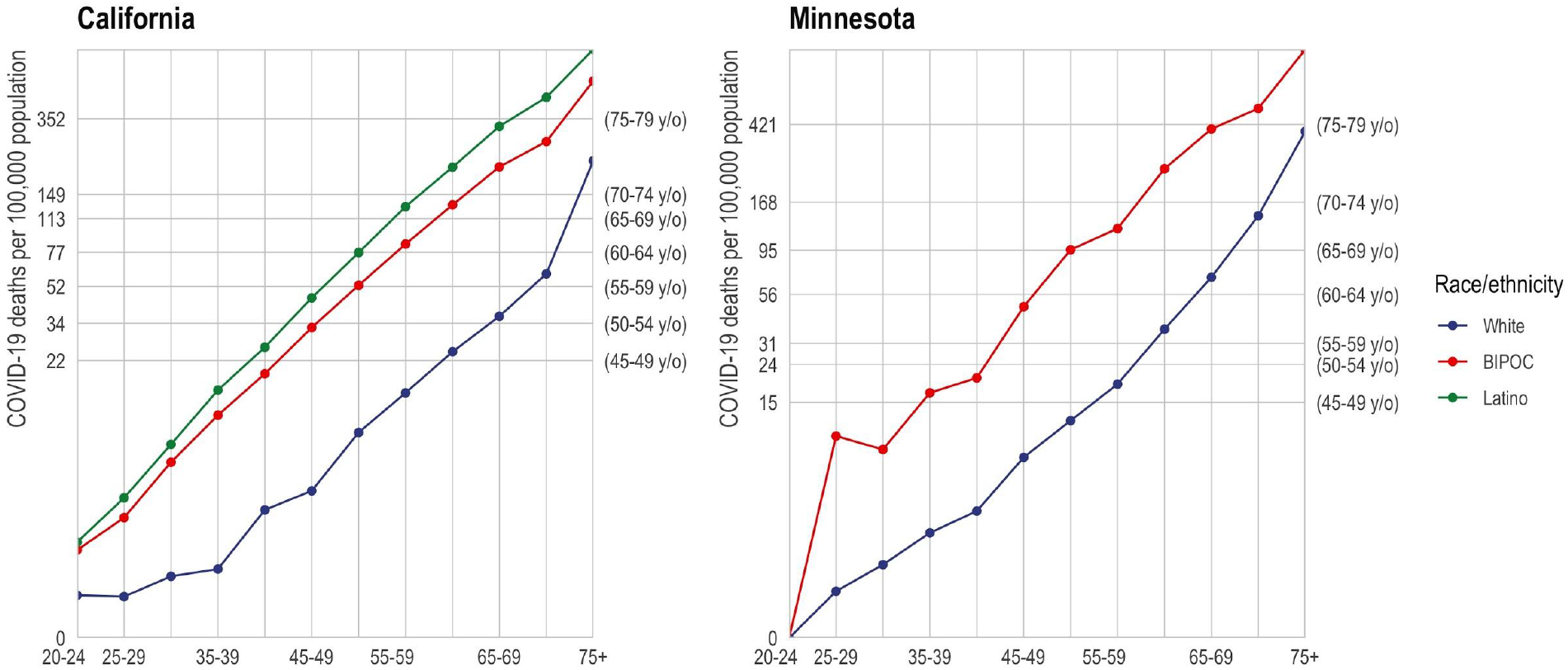
Age-specific mortality rate for white, BIPOC, and Latino race/ethnicity. The mortality rate (y-axis) by age (x-axis) varies by race/ethnicity (colors) with the non-Hispanic white population (blue) experiencing substantially lower mortality at any age relative to the BIPOC (red) and Latino (green) populations. Age-based eligibility rules ignore this variation.

**Figure S3.**
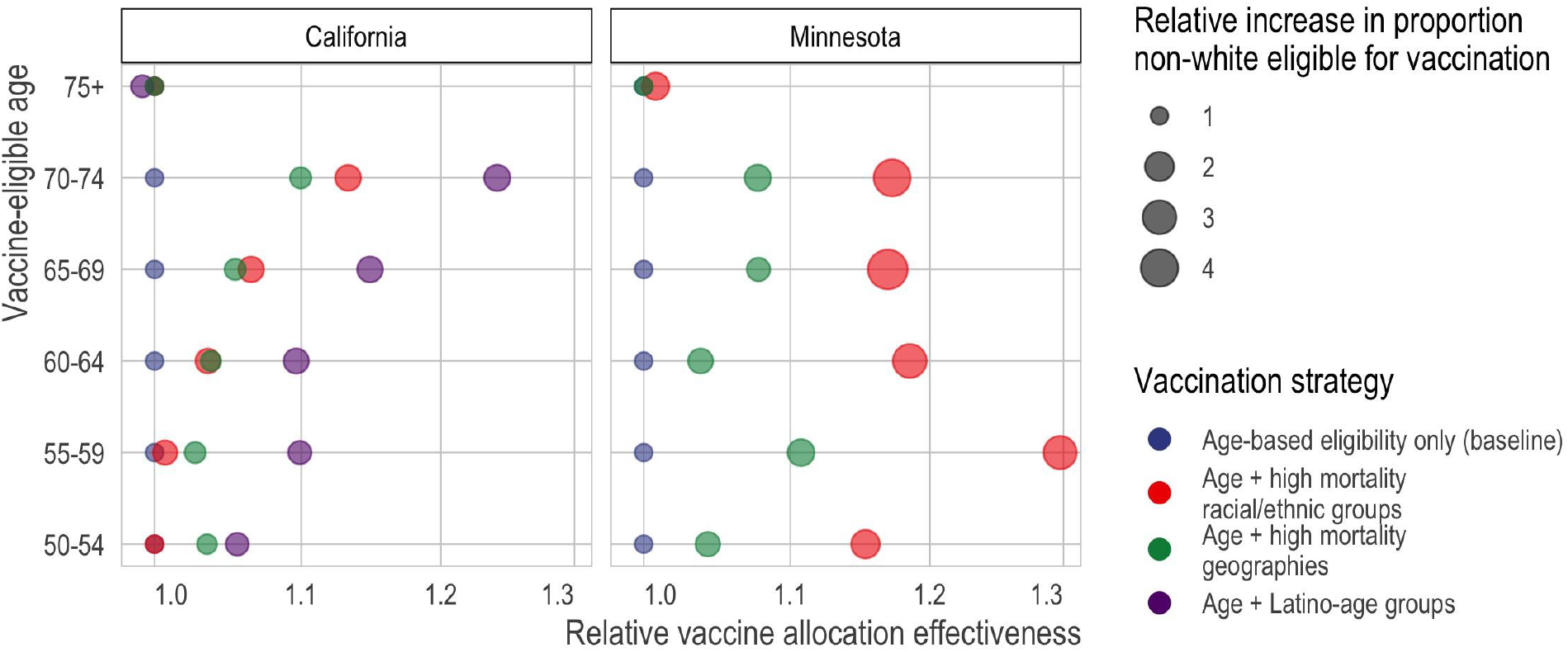
Death rate of vaccine-eligible groups under different vaccination scenarios, relative to the death rate under purely age-based eligibility. Here, we portray the same comparisons as Figure 3 in relative, rather than absolute, scale. Specifically, we compare the targeted mortality rate (x-axis) of different types of vaccine allocation strategies (color) based on age (y-axis). Specifically, we compare age alone (blue), age in combination with racial/ethnic groups (red), age in combination with high mortality locations (green), or age in combination with the highest mortality racial/ethnic group, which in California is Latinos (purple). In all cases and across all age limits, incorporating additional, younger but higher risk groups improves the efficiency of the rollout and improves racial/ethnic equity. In California, incorporating high mortality geographies (green) achieves nearly the same efficiency as incorporating high mortality racial/ethnic groups (red). In Minnesota, incorporating high mortality racial/ethnic groups always outperforms incorporating high mortality geographies; however, high mortality geography still improves the alignment of vaccine allocation with COVID-19 mortality risk.

**Figure S4.**
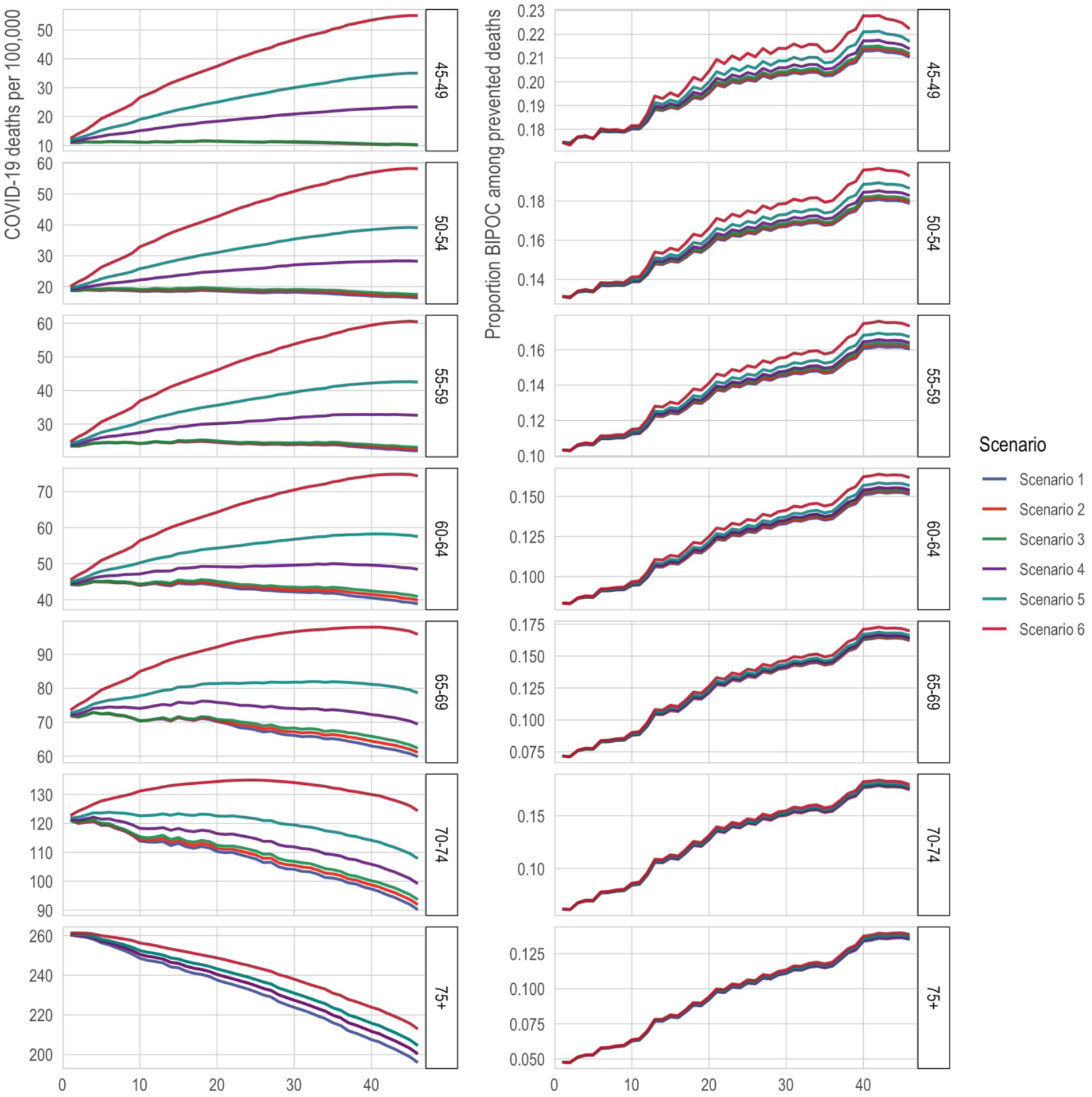
Death rates (left) and proportion nonwhite (right) among the eligible with direct targeting of high-mortality Census tracts under the alternative classification of long-term care facilities. The x-axis is the number of tracts in which all adults (ages 20+) are prioritized for vaccination. The y-axis in the left column is is 2020 COVID-19 deaths per 100,000; a higher death rate among the eligible indicates better targeting of vaccines toward high-risk individuals. The y-axis in the right column is the proportion of those eligible for vaccination who are BIPOC. The lines correspond to alternative scenarios as described in the text of the Materials and Methods. This figure reproduces the analysis in Figure 4 and Figure 5 under an alternative classification designed to produce an overcount of deaths in long-term care facilities (LTCFs), to ensure that results about prioritizing high-mortality tracts are not driven by unidentified deaths in LTCFs.

**Figure S5.**
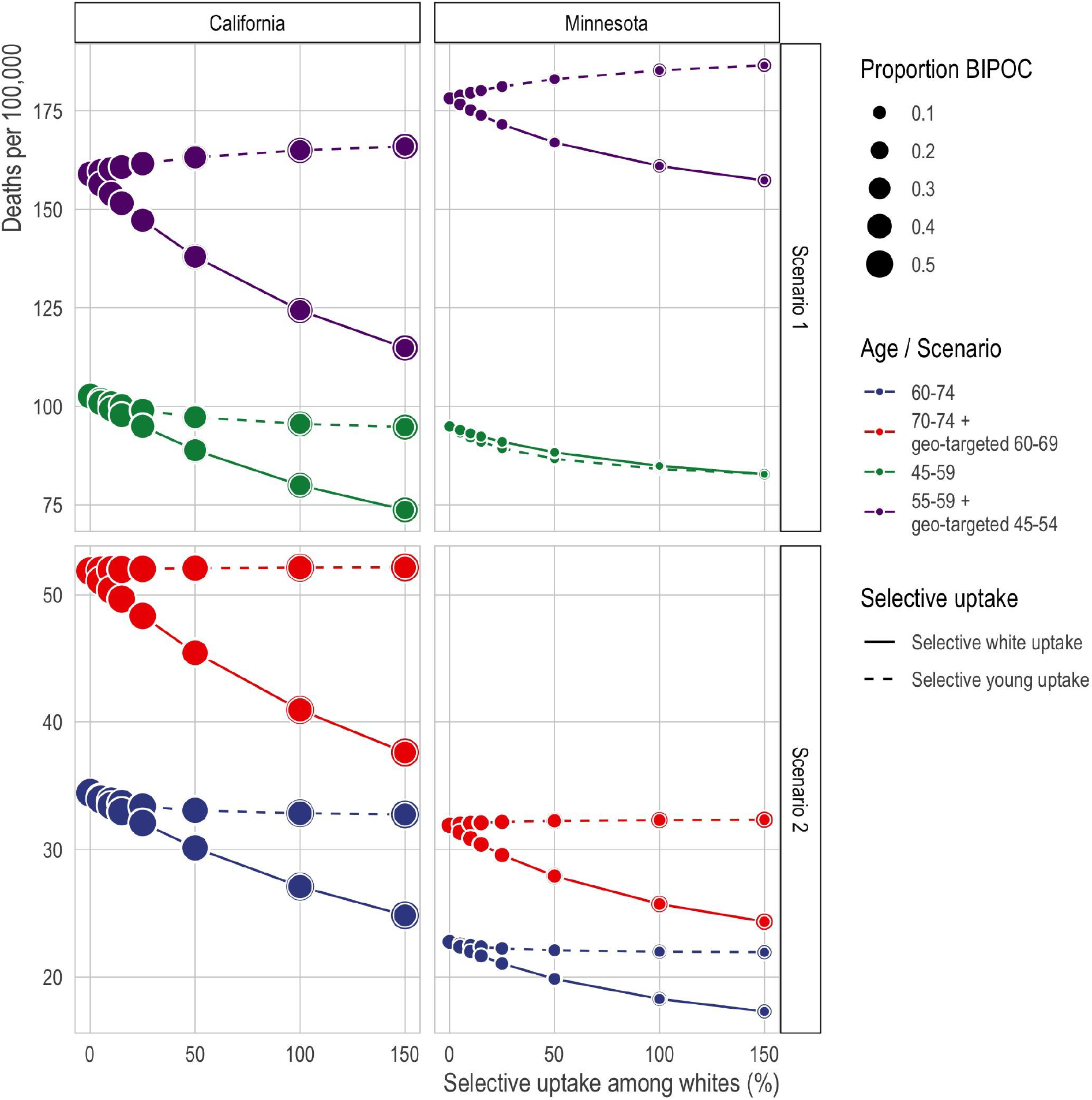
Comparison of age-alone vs. and-and-geography vaccination schedules (comparison set 4) in the presence of selective uptake by white populations. The x-axis is the percent by which vaccine uptake among the eligible is increased for white populations or for each successively younger five-year age group. The y-axis is the 2020 COVID-19 death rate per 100,000 among the eligible populations; a higher death rate among the eligible implies better targeting of vaccines toward high-risk individuals.

## Supplemental Tables

**Table S1.** Death rates and proportion white among the eligible population under various hypothetical vaccination schedules.

| State aggregate age defining eligibility | California |  | Minnesota |  |
| --- | --- | --- | --- | --- |
|  | Death rates of eligible | Percent BIPOC among eligible | Death rates of eligible | Percent BIPOC among eligible |
| 50-54, age only | 34 | 58 | 24 | 14 |
| 50-54, plus race-age groups | 34 | 58 | 27 | 27 |
| 50-54, plus Latino-age groups | 38 | 69 |  |  |
| 50-54-59, plus geog-age groups | 36 | 60 | 25 | 19 |
| 55-59, age only | 52 | 53 | 31 | 11 |
| 55-59, plus race-age groups | 52 | 71 | 40 | 32 |
| 55-59, plus Latino-age groups | 58 | 65 |  |  |
| 55-59, plus geog-age groups | 53 | 58 | 34 | 19 |
| 60-64, age only | 77 | 49 | 56 | 9 |
| 60-64, plus race-age groups | 80 | 67 | 67 | 28 |
| 60-64, plus Latino-age groups | 89 | 61 |  |  |
| 60-64, plus geog-age groups | 80 | 51 | 59 | 13 |
| 65-69, age only | 113 | 46 | 95 | 8 |
| 65-69, plus race-age groups | 121 | 67 | 111 | 37 |
| 65-69, plus Latino-age groups | 134 | 68 |  |  |
| **65-69, plus geog-age groups** | 119 | 50 | 102 | 10 |
| **70-74, age only** | 149 | 42 | 168 | 7 |
| **70-74, plus race-age groups** | 169 | 63 | 197 | 26 |
| **70-74, plus Latino-age groups** | 190 | 65 |  |  |
| **70-74, plus geog-age groups** | 164 | 46 | 181 | 11 |
| **75+, age only** | 352 | 41 | 421 | 5 |
| **75+, plus race-age groups** | 352 | 41 | 424 | 9 |
| **75+, plus Latino-age groups** | 362 | 48 |  |  |
| **75+, plus geog-age groups** | 352 | 41 | 421 | 5 |

